# Investigation of intra-subtype recombination in Zimbabwean HIV 1 subtype C using bioinformatics. (December 2019)

**DOI:** 10.1101/2022.10.29.22281702

**Authors:** Milton S Kambarami, Manasa Justen

## Abstract

*Intra-subtype* recombination is a common phenomenon among HIV strains which involves transfer of genetic information between two viral strains belonging to the same HIV subtype. However, more research has been done on *inter-subtype* recombination which is the transfer of genetic information between two strains belonging to different HIV subtypes. In this study HIV 1 subtype C *env* sequences from Zimbabwean nucleotide sequences (n=2 915) downloaded from Los Alamos HIV databases were used to investigate intra-subtype recombination. Using the GENECUTTER tool, ∼44.4% of whole genomes were selected to have informative *env* regions (n = 1 295). Clustal was used for Multiple Sequence Alignment and RDP 4 was used for analysis and detection of Intra-subtype recombination. Apparently, GENECONV included in RDP 4 detected 47 recombination events. One of the most informative recombination events was further analysed using UPGMA phylogenetic tree to understand the extent of the intra-subtype recombination.

## Introduction

Genetic recombination is transfer of genetic information from one sequence to another. In cases where the evolutionary origin of the shared nucleotide sequences is the same, it is termed homologous recombination. Recombination between nucleotide sequences which do not belong to the same evolutionary origin is called non-homologous recombination (Cromie *et al. 2001*).

Recombination among HIV strains can be sub-categorised into inter-subtype recombination and intra-subtype recombination. Inter-subtype recombination involves transfer of genetic information between two HIV sequences which belong to different HIV subtypes, these recombinants are usually termed CRFs (Circulating Recombinant Forms). Intra-subtype recombination involves transfer of genetic information between two HIV sequences which belong to the same HIV subtype. HIV sequences which have undergone intra-subtype recombination tend to have high sequence similarity compared to sequences which have undergone inter-subtype recombination, this makes intra-sutype recombination very difficult to detect than inter-subtype recombination

Recombination events play a significant role in evolution and diversity of Human Immunodeficiency Virus (HIV). Statistically, HIV can adapt faster if mutations happen in a number of viral copies simultaneously than when mutations happen in one viral copy per given time. Recombination plays an important role in ensuring transfer of these advantageous traits from one virus to another as well as elimination of ineffecient traits thus resulting in a fit virus which can adapt in harsh conditions and a weaker virus which is waded off by environmental pressures.

It has been observed that recombination between different gene segments of HIV make an important contribution to diversity in the *env* gene (Charpentier *et al*. 2005). The *env* gene codes for the envelope which surrounds HIV, this envelope changes for the virus to evade the host immune system. These changes are chiefly mediated by genetic recombination and polymorphism mutations.

According to Schierup and Hein, 2000, Posada and Crandall, 2002, the strength of a recombination signal when using recombination detection and analysis methods is governed by: (i) the number of variable nucleotides different in both parents and (ii) the proportion of variable nucleotides contributed by each parent. Computationally, it is an np-hard task to find the true extent of intra-subtype recombination.

Failure to recognise recombination effects can account for compromised validity of currently available computational tools for studying molecular evolution. (Martin, Lemey and Posada, 2011). Recombination events can distort phylogenetic signals such that in phylogenetic analyses, variant sites in recombinant portions of the genomes appear as homoplasy instead of a single lineage. This results in longer branch lengths and overestimation of the substitution rate heterogeneity (Liu *et al*. 2004, Schrieup and Hein, 2000).

In this study we set out to investigate intra-subtype recombination in Zimbabwean HIV sequences downloaded from Los Alamos HIV database using bioinformatics tools.

## Methodology

All of the Zimbabwean HIV sequences were downloaded from the Los Alamos HIV database which had a size of ∼16 Megabytes (MB) and n= 2915 (number of sequences). Only HIV 1 subtype C sequences with the *env* gene were selected manually using Mega X to reduce the size of the dataset so it could be inserted into Los Alamos *GeneCutter* tool with a limit of <10MB insertion data. The *GeneCutter* tool selects specified regions in a given dataset in this case the *env* gene, the result had n= 1295 informative sequences and a data size of ∼4.2 MB.

Clustal which is included in the Mega X software was used for Multiple Sequence Alignment. The offline Clustal was favourable for the task compared to the online Clustal alignment tool because the maximum data upload size online is 4MB and no sequence could be compromised to not participate in analyses that would follow. The Multiple Sequence Alignment process was left to run overnight until complete. The resultant alignment file was saved in FASTA format.

The alignment FASTA file was uploaded onto Recombination Detection Program 4 (RDP4) for detection and analyses of intra-subtype recombination. The GENECONV recombination method registered a remarkable number of recombination signals compared to other recombination detection signals in RDP4. One of the recombination event (number 35) showed significant intra-subtype recombination event, hence more phylogenetic tests were carried out to ascertain the precision and accuracy of the obtained result shown in Fig 1.

**Fig 1:**
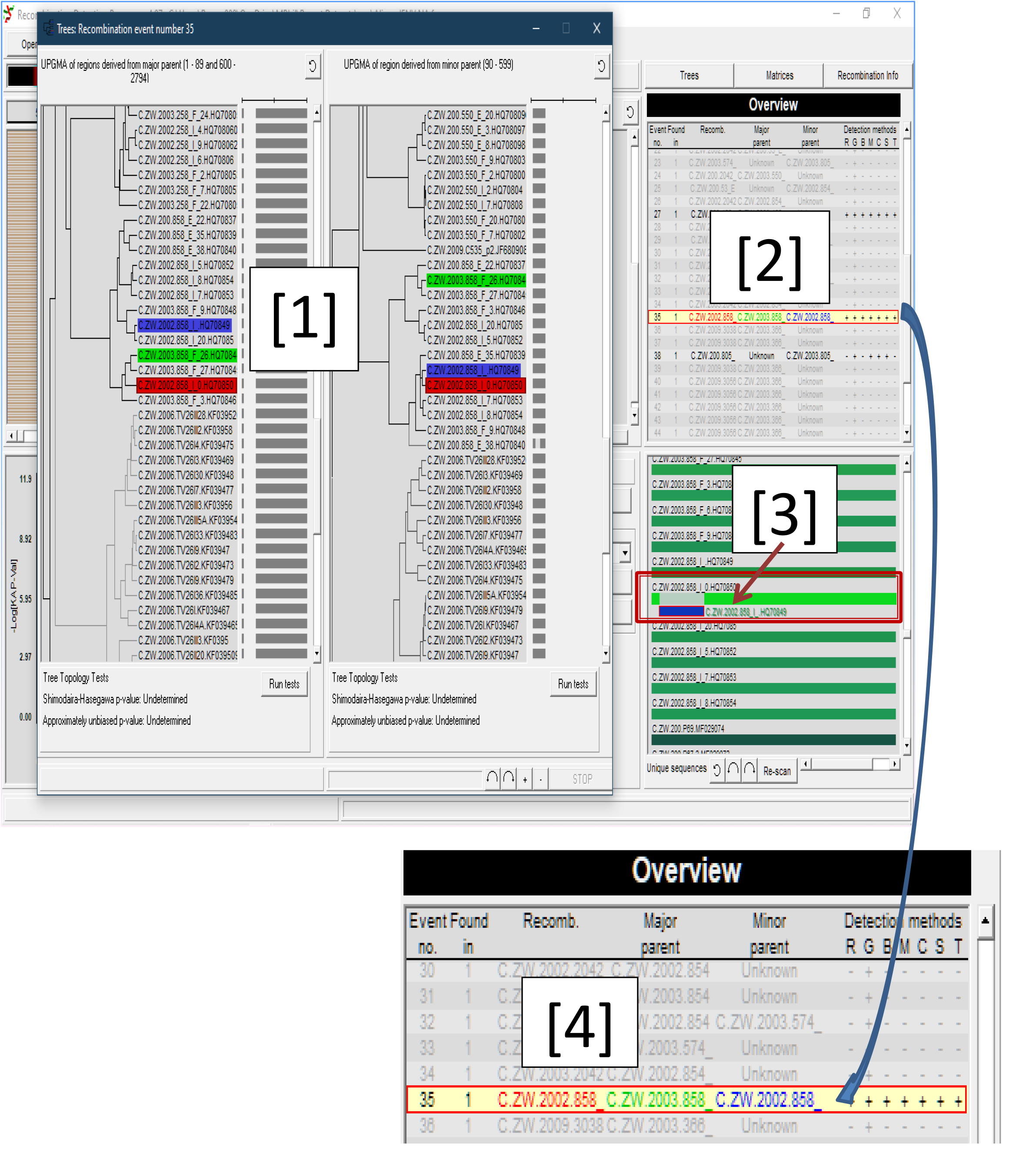
Overall analysis of recombination using GENECONV analysis method in RDP 4. The sequences are colour-coded as follows: Recombinant = Red, Major Parent = Green, Minor Parent = Blue. [1] UPGMA Phylogenetic tree to show how the best representative of evidence of recombination is related, the left phylogenetic tree shows the related region derived from the Major Parent (The derived regions’ nucleotide positions provided) in contrast to the minor parent, the right phylogenetic tree illustrates the phylogenetic relationship of the region derived from the minor parent and the recombinant in contrast to the major parent (the derived region’s nucleotide positions provided). [2] General Overview of recombination events detected by the primary recombination detection methods which are available in RDP 4. [2] is zoomed into [4] to show a visible illustration of recombination event 35, [-] and [+] signs show whether a recombination event was not detected or detected respectively by a recombination method represented by the its first letter on top. [3] is a schematic diagram to show regions of recombination in the sequences of the recombinant and the minor parent.

## Results

## Discussion

According to overall observed results in Fig 1, for a given population of viral copies in a T cell there is ∼4% chance that the Zimbabwean HIV 1 subtype C virus undergoes recombination. This was observed using the GENECONV recombination detection method. Such high rate of intra-subtype recombination has an impact on virus evolution and diversity. Recombination event number 35 was selected for further study because the contributing sequences had ‘known’ names. Furthermore, the recombination event was detected by all detection methods included in RDP 4 shown in Fig 1 frame 4. Such a result supports a very high possibility that there was an intra-subtype recombination between selected sequences.

Intra-subtype recombination has an impact on viral evolution and viral diversity. Evolution of HIV has been the chief drive of anti-retroviral drug resistance and mode of action of HIV to evade the host-immune system. Targeting genes which undergo slower recombination/evolutionary rates then designing a drug with high affinity for the selected gene using molecular docking techniques, such a drug can combat the proliferation of HIV in the host.

The *env* gene was chosen for this study because it is very susceptible to genetic changes which include recombination events and polymorphisms (Charpentier *et al*. 2005). These changes are necessary for the virus to evade the host immune system and anti-retroviral drug effects.

Intra-subtype recombination studies of HIV type 1 subtype C are significant in Zimbabwe because it is the most prevalent HIV strain in Zimbabwe with sequences uploaded on the Los Alamos database amounting to 98.4% of Zimbabwean HIV sequences. With this information obtained through bioinformatics, it will be easier to design drugs that halt intra-subtype recombination followed by eradication of the weaker HIV copies thus lowering HIV burden in the host cell.

Simian Immunodeficiency virus (SIV) has a close phylogenetic relationship with HIV, SIV crossed species barrier to infect Humans as an adaptation to new host (Williams and Burdo, 2009). Recombination dynamics of zoonotic viruses can aid in predicting when these virus will adapt to infect a related species such that vaccines would have been designed before or on the onset of zoonotic infection.

## Data Availability

All data used in present study are available upon reasonable request to the authors.

